# Predicting Motor Vigor from EEG Phase Amplitude Coupling (PAC) in Parkinson’s Disease: Effects of Dopaminergic Medication and Non-invasive Modulation

**DOI:** 10.1101/2024.02.20.24303077

**Authors:** Alireza Kazemi, Salar Nouri, Maryam S. Mirian, Soojin Lee, Martin J. McKeown

**Affiliations:** Center for Mind and Brain, Department of Psychology, University of California, Davis, Davis, CA, United States; School of Electrical and Computer Engineering, College of Engineering, University of Tehran, Tehran 14395-515, Iran; Pacific Parkinson’s Research Centre, Djavad Mowafaghian Centre for Brain Health, University of British Columbia, Vancouver, BC V6T 2B5, Canada; Faculty of Medicine (Neurology), the University of British Columbia, Vancouver, BC V6T 2B5, Canada

**Keywords:** Parkinson’s disease, Phase-amplitude coupling, Deep neural network, Transfer learning, Motor vigor, EEG regression, Neurophysiological analysis, biomarker, brain stimulation

## Abstract

Impaired motor vigor is a defining characteristic of Parkinson’s disease (PD), yet the underlying brain mechanisms governing motor vigor (MV) remain unclear. Recent studies have suggested beta-gamma Phase-Amplitude Coupling (PAC) derived from the resting-state electroencephalogram (EEG) is a potential biomarker for PD that is modulated by Deep Brain Stimulation (DBS) and L-dopa treatment. Specifically, PAC has been suggested to be a marker of transitions between motor movements, as opposed to encoding the vigor of the current movement. Here, we comprehensively investigate the potential of various PAC interactions—across different frequency pairs—beyond the linear approaches typically employed to predict MV during motor tasks in PD and study the effects of dopaminergic medication and non-invasive Galvanic Vestibular Stimulation (GVS). We recorded EEG data from 20 PD patients and 22 healthy controls executing a simple, overlearned handgrip task. Subjects were tested on and off L-dopa medication and with and without GVS (multi-sine either 50-100Hz, 100-150Hz). In a preliminary linear (LASSO-based) analysis comparing various PACs and a broad range of commonly used EEG features, PAC features were found to be crucial for predicting MV approximately equally in PD and HC. Initial findings from the linear analysis showed PAC as a significant indicator for MV in both groups, although with variability in cross-validation that implied a complex, non-linear relationship between PAC and MV. To extensively investigate the PAC-MV relation, we used a deep convolutional neural network (PACNET)—developed based on pre-trained VGG-16 architecture—to estimate MV from PAC values. In both PD and HCs, delta-beta, theta-, alpha-, and beta-gamma PACs were important for MV prediction. In PD subjects, GVS affected delta-beta, theta-gamma-, and beta-gamma PACs role in MV prediction, which was sensitive to different GVS stimulation parameters. These PACs were also relevant for PD patients’ MV prediction after L-dopa medication. This study supports the hypothesis that EEG PAC across multiple frequency pairs, not just beta-gamma, predicts MV and not just motor transitions and can be a biomarker for assessing the impact of electrical stimulation and dopaminergic medication in PD. Our results suggest that PAC is involved in MV, in addition to a range of previously reported cognitive processes, including working and long-term memory, attention, language, and fluid intelligence. Non-linear approaches appear important for examining EEG PAC and behavior relations.

## Introduction

Motor vigor (MV) represents a crucial aspect of the motor system, potentially impacted by various diseases. It serves as an indicator of an individual’s capacity to utilize physical energy effectively and efficiently in executing motor tasks (Shadmehr & Ahmed, 2020). MV can be modulated by an interplay of neural, physiological, psychological, and environmental influences. The basal ganglia appear vital for weighing the effort/reward ratio for performing certain motor tasks, but cortical involvement is likely also significant (Uehara et al., 2023). This involvement implies that EEG, which primarily measures cortical activity (Constant & Sabourdin, 2012), could be an effective tool for monitoring MV.

In the context of Parkinson’s disease (PD), MV gains particular significance. One of PD’s hallmark symptoms, bradykinesia—the marked slowness of movement—is essentially a manifestation of diminished MV. The full underlying brain mechanisms governing impaired vigor are not completely understood, but PD bradykinesia has been linked to changes in beta-band (13– 30 Hz) Local Field Potentials (LFPs) within the basal ganglia (Little et al., 2012). LFPs tend to be more sensitive than the EEG, but the scalp EEG has also previously been shown to be abnormal in PD, with beta-band oscillations over sensorimotor electrodes in PD patients off medication having altered morphology (Jackson et al., 2019) and machine learning approaches being able to predict overall disease severity (Arasteh et al., 2021). Our prior research has shown that standard features derived from EEG data can accurately predict MV in both healthy individuals and those with PD (Kazemi et al., 2021). In this study, we further investigate the connection between Motor Vigor (MV) and EEG features.

One feature of abnormal brain oscillations in PD is altered Phase Amplitude Coupling (PAC). PAC calculates the relations between the phase of low-frequency oscillations and the amplitude of high-frequency oscillations. Under normal conditions, PAC can be modulated by visual stimuli (Voytek et al., 2010) and motor imagery (Gwon & Ahn, 2021). In PD subjects, 13-30 Hz (phase) and 200-500 Hz (amplitude) PAC in the LFPs from the subthalamic nucleus (STN) predict response to Deep Brain Stimulation therapy (Yang et al., 2014). In an animal model, delta-high-gamma PAC in the motor cortex and striatum has been observed by dopamine D2 receptor blockade (Reakkamnuan et al., 2023). Altered PAC in PD has been observed at rest in both the MEG (Mertiens et al., 2023) and EEG (Miller et al., 2019; Zhang et al., 2021). This has led to speculation that PAC might be a valuable biomarker to target neuromodulation strategies (Hwang et al., 2020; Salimpour et al., 2022; Yeh et al., 2023). Altered PAC may also be important for freezing of gait in PD (Karimi et al., 2022).

PAC has been implicated in several cognitive processes (Sacks et al., 2021), likely because PAC may facilitate communication between spatially distributed cortical networks (Turi et al., 2020). Theta–gamma PAC is important for visual and auditory working memory (Axmacher et al., 2010; Kaminski et al., 2019) and encoding and retrieval in long-term memory (Friese et al., 2013; Köster et al., 2019; Lara et al., 2018). PAC has been associated with verb generation (Doesburg et al., 2012), language prediction (Wang et al., 2018), selective attention (Doesburg et al., 2012; Saalmann et al., 2012) and possibly fluid intelligence (Pahor & Jaušovec, 2014). Interestingly, PAC may be important for non-invasive brain stimulation techniques. After a 2-week course (10 sessions) of repetitive transcranial magnetic stimulation (rTMS) over the dorsolateral prefrontal cortex to treat depression, PAC was increased at rest. Theta–gamma transcranial alternating current stimulation (tACS) has a greater effect on working memory than theta tACS alone (Alekseichuk et al., 2016).

While most studies examining altered PAC in PD have focused on rest, a recent study demonstrated that beta-gamma PAC was altered during transitions between different movement states, with the speculation that this was related to bradykinesia in PD (Gong et al., 2022). Here, we explore an alternate hypothesis, namely that PAC, and not just beta-gamma PAC, is directly associated with MV, as opposed to movement transitions. We examine both linear and nonlinear approaches to determine if PAC can accurately predict MV, as exhibited in a simple, overlearned hand-squeeze task, in PD subjects and controls. We show that EEG-derived PACs at multiple frequencies are important features for predicting MV.

## Materials and Methods

### Participants

All participants provided written, informed consent prior to participating. The research protocol was approved by the Clinical Research Ethics Board at the University of British Columbia. Participants included 20 healthy controls (HC) and 20 Parkinson’s disease (PD). Data from PD patients were collected under two medication conditions: Off (after > 12hr withdrawal of dopaminergic medication: PD-off) and On (after taking their regular medication: PD-on). Table 1 presents the demographic and clinical characteristics of participants included in this study. For further details regarding inclusion and exclusion criteria, please check (Lee, 2019; Lee et al., 2019).

**Table 1.** Demographic and clinical characteristics of the participants.

|  | PD (N=20) | HC (N=20) |
| --- | --- | --- |
| Age (year) | 67.0 ± 7.0 | 467.5 ± 6.4 |
| Sex (male/female) | 10/10 | 10/10 |
| Disease duration (year) | 7.6 ± 4.3 | - |
| UPDRS III | 23.5 ± 9.8 | - |
| UPDRS III item 3.16 (kinetic tremor) | 2.3 ± 1.3 | - |
| UPDRS III item 3.17 (rest tremor) | 2.4 ± 2.2 | - |
| Hoehn and Yahr scale | 1 - 2 | - |
| Levodopa Equivalent Daily Dose (mg) | 708.2 ± 389.9 | - |

### Behavioral Data Collection

The experimental paradigm was a simple visuomotor task conducted in different blocks of 10 trials. In each trial, participants were given a visual fixation for a brief jittered time (1500ms + random delay of 0-500ms, uniformly distributed), followed by a “Go” signal. They were instructed to squeeze the bulb as quickly and strongly as possible when they saw the “Go” signal on the screen (See Figure 1.A).

**Figure 1.**
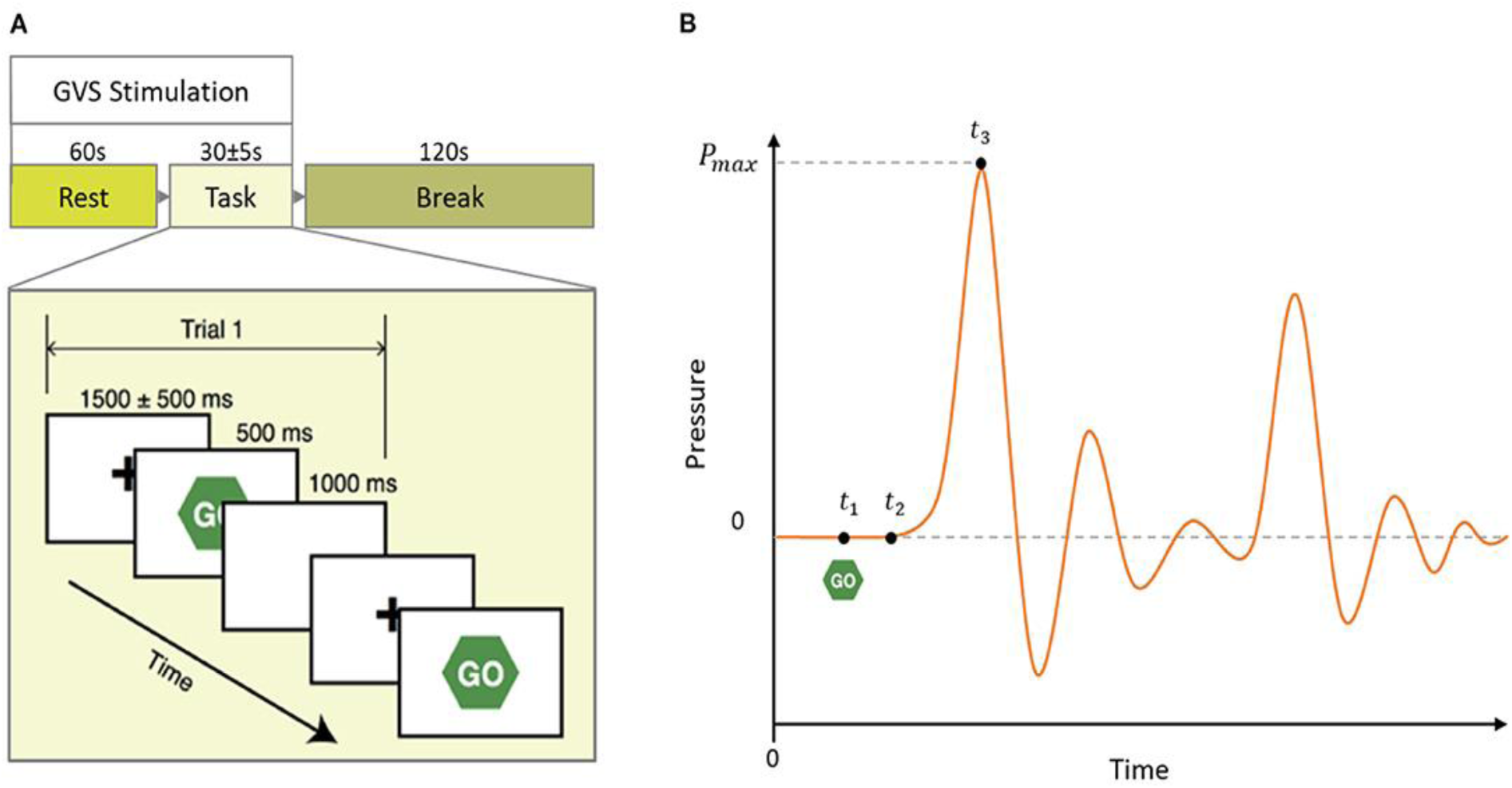
A-Schematic of a block of the experimental paradigm in which 60 seconds of rest is followed by 10 task trials and 120 seconds of break time. In each block, GVS stimulation (Sham, GVS1: 50–100 Hz, and GVS2: 100–150 Hz) was delivered during the rest and task period. (B) Mock pressure signal of a squeezing bulb. The GO screen appeared at t_1_; the participant started to squeeze the bulb at t_2_ and reached maximum pressure at t_3_. Peak time is defined as t_3_ − t_1_. Some reduced oscillations subsequently occur because of the compliance of the squeeze bulb apparatus

Each subject’s behavioral data, including response time (RT) and peak time (PT), were recorded. Peak Time is defined as the time it takes for participants to squeeze the bulb to their maximum pressure, *t_3_ − t_1_* (See Figure 1.B). MV was estimated as the inverse of the Peak Time. Note that this includes both reaction time (*t_2_-t_1_*) and movement time (*t_3_-t_2_*).

### EEG Recording and Preprocessing

Using a 32-channel EEG cap with electrodes positioned according to the global 10-20 placement standard and two additional pairs of surface electrodes to monitor vertical and horizontal visual stimuli, EEG data were gathered from 27 scalp electrodes. Different preprocessing methods, including independent component analysis and filtering, were used for artifact rejection as described in (Lee et al., 2019). Two-way finite impulse response (FIR) filters were used to filter the preprocessed EEG signals into the standard EEG frequency bands. Further preprocessing methods are identical to (Lee et al., 2019, 2021).

### Analysis Methods

#### Preliminary Analysis

We first predicted MV from PACs using conventional linear regression optimized by the LASSO (Tibshirani, 1996) algorithm. We took our previous model (Kazemi et al., 2021) as the baseline model, which was a LASSO algorithm performed on commonly used EEG features (will be referred to as conventional EEG features) computed based on standard EEG subbands (See (Kazemi et al., 2021) for details). We then extended the prior approach by using the LASSO algorithm to predict MV when only PAC features were available and when both conventional and PAC features were available. These results were compared against our previous model which used only conventional features of EEG (Kazemi et al., 2021). Since this step was only a preliminary analysis, we only analyzed MV in the HC and PD groups off medication and without stimulation.

#### PACNET

To comprehensively examine PAC-MV relations across all sub-bands, we utilized a pre-trained deep CNN-based model with transfer learning, which we called PACNET which enabled us to extensively examine the potential complex dynamics between PAC and MV across different conditions of health status (i.e., HC and PD), stimulation (i.e., Sham, GVS1 and GVS2), and L-dopa medication (i.e., on and off). The complete pipeline of analysis carried out is shown in Figure 2. Initially, the PAC value (See supplementary Material section 2) for each pair of the five standard EEG subbands—Delta (0.5-4 Hz), Theta (4-8 Hz), Alpha (8-16 Hz), Beta (16-32 Hz), and Gamma (32-45 Hz)—was calculated to create a 2-dimensional representation of PAC values using TensorPAC (Combrisson et al., 2020), an open-source Python library for PAC investigation of neurophysiological data. Subsequently, we used the VGG-16 architecture (Simonyan & Zisserman, 2015) for transfer learning (called PACNET) to train a model to predict MV. TensorPAC images were created with dimensions 32 * 64 * 3 (due to VGG16 structure) as PACNET inputs. At each training iteration, the initial weights of the model were set to the values of a pre-trained model on the ImageNet dataset, which contains over 14 million images belonging to 1000 classes (Simonyan & Zisserman, 2015). The architecture of PACNET, based on the VGG-16 architecture, is described in detail in the Supplementary Material section 3. A single model was trained on all conditions (i.e., Health, Stimulation, and Medication). We subsequently evaluated the performance of the trained model to be comparable between the conditions (See supplementary Material section 4 for performance metrics to evaluate the training). The baseline performance of the model in predicting MV using PACs was compared with the linear regression models utilized in the preliminary analysis.

**Figure 2.**
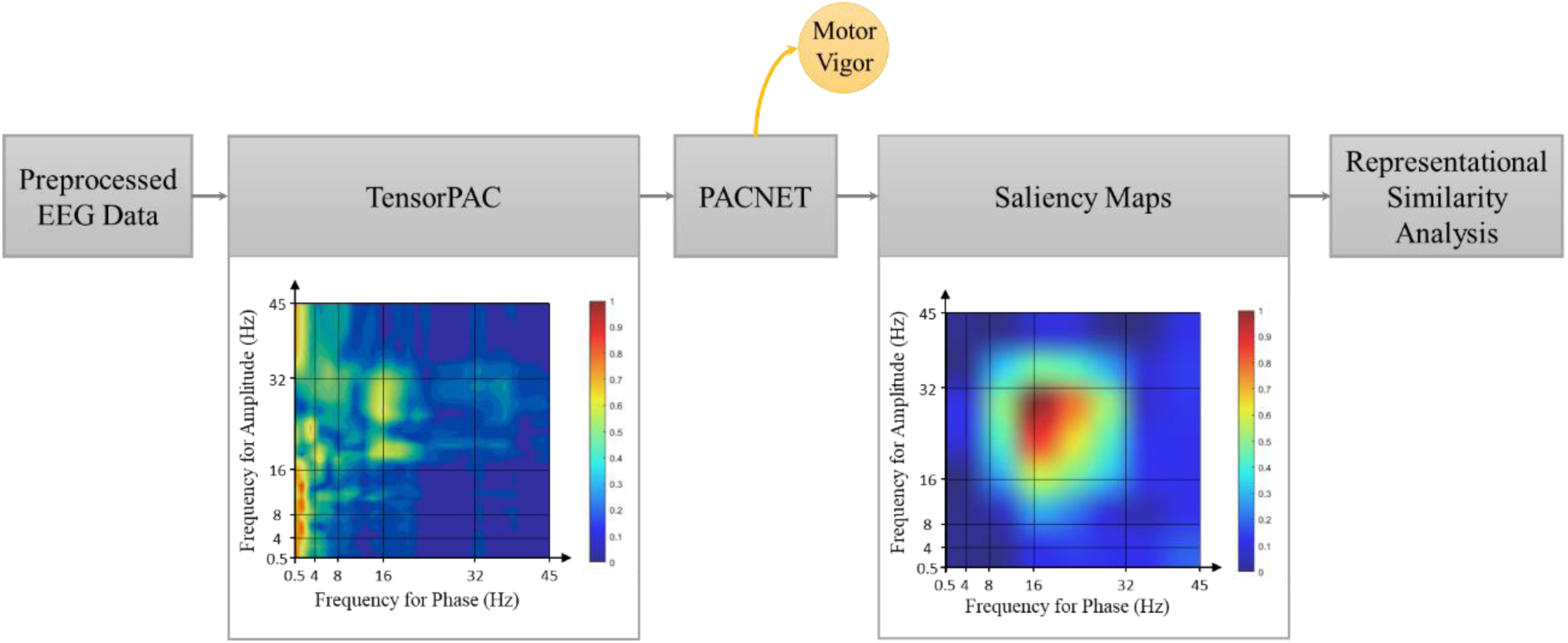
Schematic view of the proposed approach. A more detailed representation of the pipeline can be found in the Supplementary Material Section 1.

The convolutional layers within PACNET are adept at identifying and learning patterns that span the complete frequency range depicted in tensor PAC images. This capability enables them to capture local and global features, essential for a nuanced understanding of the complex interactions in neural signals. Consequently, we hypothesized that our model could conduct an exhaustive analysis of the spectral characteristics inherent in PACs. This approach is advantageous as it avoids the limitations of focusing solely on discrete frequency bins, potentially uncovering aspects that might otherwise be overlooked.

We employed bootstrapping technique (Kazemi et al., 2021) to train PACNET and evaluate its saliency maps, which consisted of 40 separate iterations of training and testing. At each iteration, 80% of randomly selected trials were used for model fitting (i.e., training PACNET or fitting Lasso) to predict MV, and the remaining 20% of trials were used to estimate the performance of the trained model in predicting MV.

#### PAC-MV Relation Analysis

We used a standard method to quantify the informativeness of the inputs by examining the last convolutional layer of the trained model (Alqaraawi et al., 2020; Gao et al., 2023). We employed Gradient-weighted Class Activation Mapping (Grad-CAM) (Simonyan et al., 2013) to highlight the influential critical PACs in MV estimation. The gradients of each TensorPAC image entering the PACNET’s final convolution layer (i.e., saliency map) identify the importance of different PACs for estimating MV (See Supp Mat section 5 for more details).

To examine the consistency of the patterns in saliency maps across different boot-strapping runs, we utilized representational similarity analysis (RSA) (Kiat et al., 2022; Kriegeskorte, 2008). The degree to which PACs consistently demonstrated informativeness for MV prediction was estimated through pairwise Pearson’s correlation tests between regions of interest in the saliency maps. The regions of interest were defined to cover the areas that are corresponding to standard PAC frequency pairs (see Figure 2B). Higher similarity scores corresponded to greater consistency in the informativeness of a PAC across images (i.e., these regions were more frequently identified as informative by the PACNET during the MV prediction process). However, to eliminate the effect of areas of the saliency maps that were highly similar because of not being informative—having low values—we only included PAC regions in which their mean and median were higher than the mean and median of the whole saliency map. Additionally, we controlled all the similarity scores based on the variability in the TensorPAC images. Since we were looking for consistent and cross-condition informativeness, we reasoned that PACs with less variability in the TensorPAC images and large variability in the saliency maps are more likely captured by the PACNET to account for variabilities between trials of the same condition rather than consistent effects of conditions across trials. Consequently, we excluded similarity scores for subregions with statistically less variability in the TensorPAC images compared to the same region in the saliency maps based on a one-sample t-test on the difference between their similarity scores.

## Results

### Behavioral Performance

We used a mixed ANOVA model (See Figure 3) to compare MV between HC and PD across different stimulations. We found a main effect of stimulation, *F*_2, 66_ = 50.98, *p* < 0.001, such that the average MV in the sham condition was significantly lower (*M* = 1.39e-3, *SD* = 1.74e-4) than the average MV in both GVS1 (*M* = 1.54e-3, *SD* = 1.93e-4), *t*(34) = 7.21, *p* = 0.001, and GVS2 (*M* = 1.54e-3, *SD* = 1.94e-4), *t*(34) = 7.14, *p* < 0.001. We found no other main or interaction effects (*ps* > 0.156).

**Figure 3.**
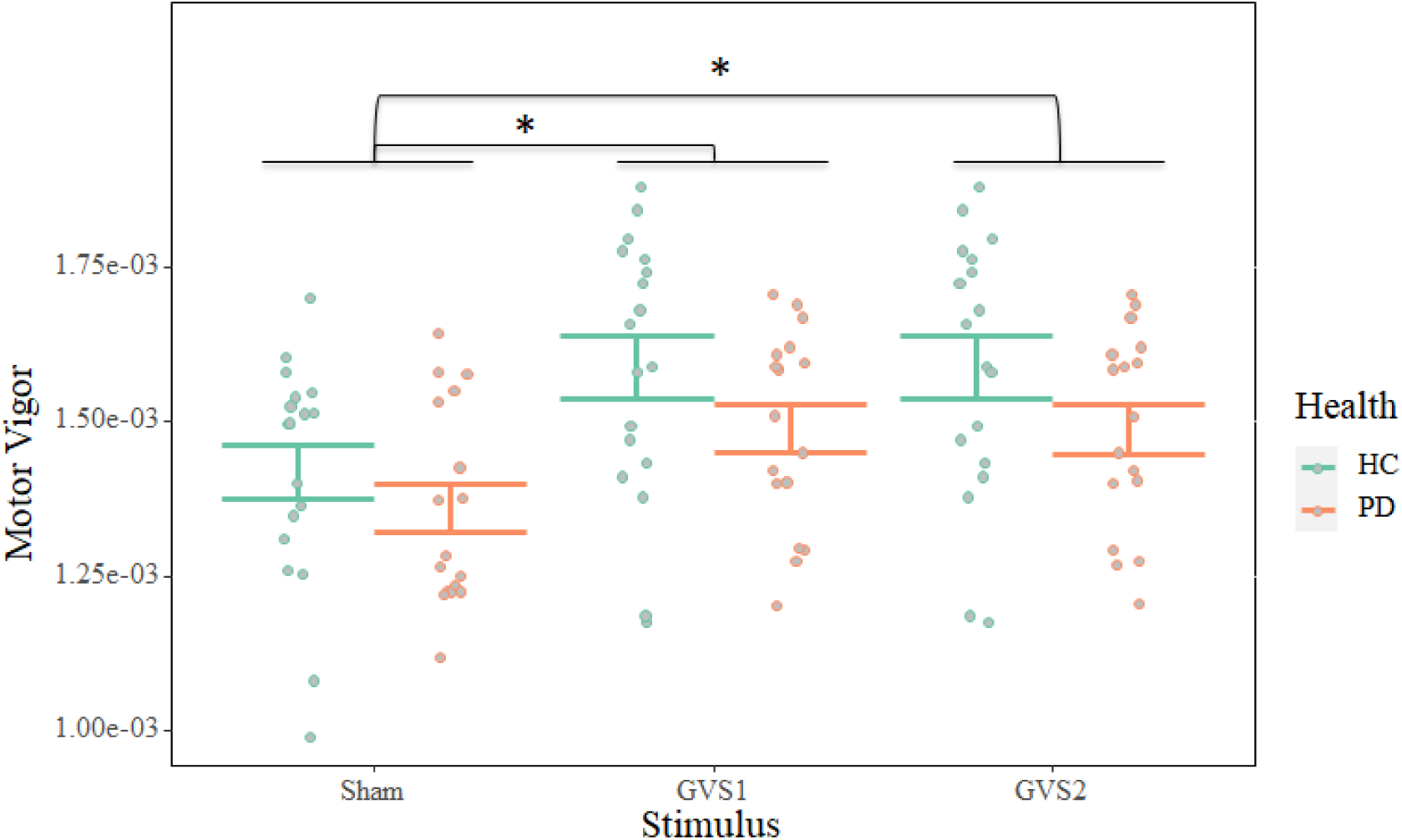
Motor Vigor distribution for each condition. *indicates that the Bonferroni corrected p-value was significant (<0.05). Error bars indicate the standard error around the mean.

### Preliminary Analysis and PACNET Baseline Performance

PACNET successfully trained on the training sets in all 40 iterations (details of the training metrics are provided in the Supplementary Material, Section 6). The performance (based on the correlation between actual MV and the predicted MV) of the LASSO linear regression on predicting MV from different feature subsets (only PAC features, only conventional EEG features, and both) along with PACNET’s performance is shown in Figure 4.

**Figure 4.**
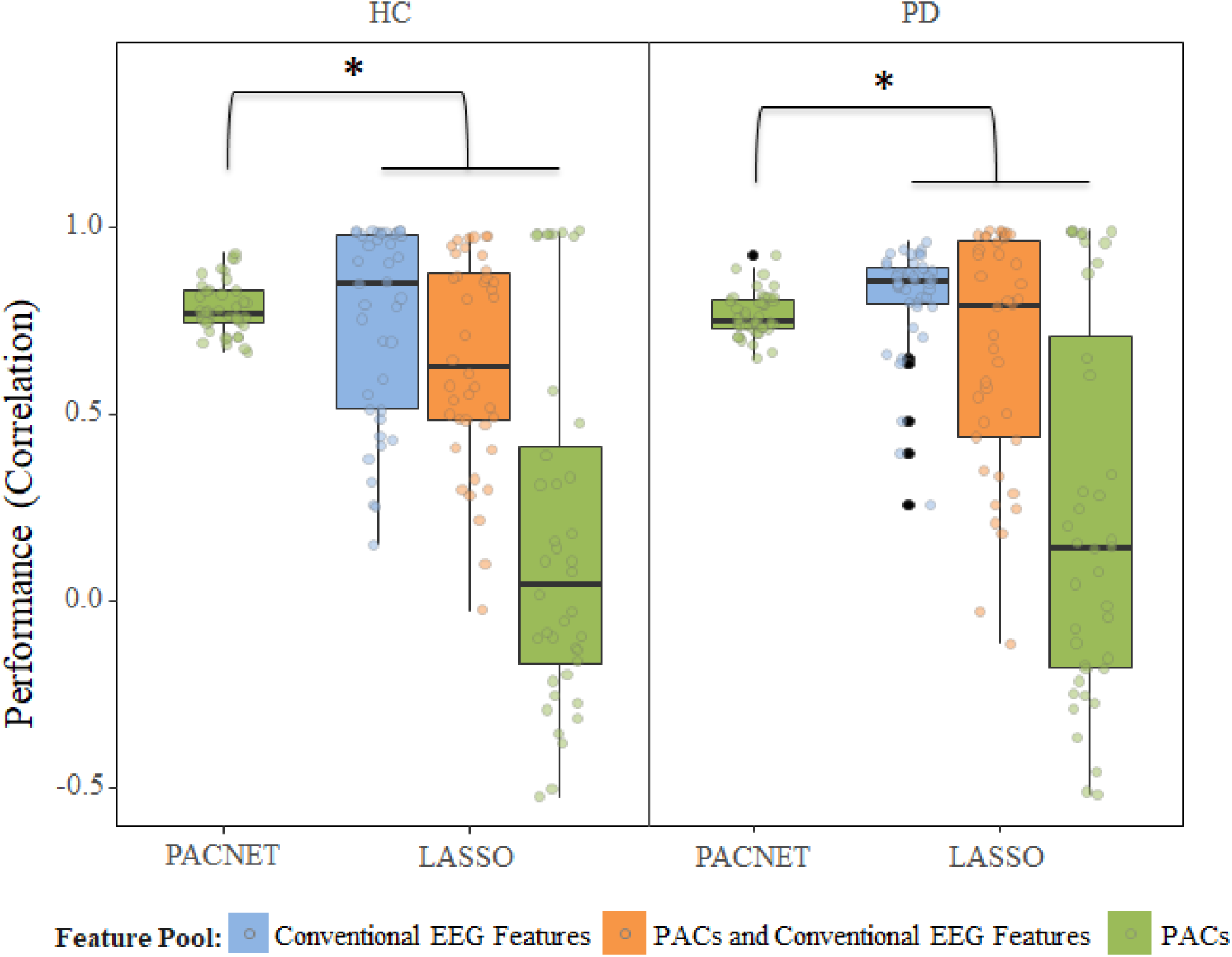
Comparing the predicted vs. actual MV for PACNET and LASSO with different feature pools of conventional EEG features, PAC features, and both. *indicates that the Bonferroni corrected p-value was significant (<0.05).

The LASSO algorithm on only PAC features did not converge to one model in more than 50% of our boot-strapped simulation runs and exceeded the maximum number of iterations (*N* = 1000). Comparing the variance of model performance in different iterations between models using an F-test demonstrated that PACNET had the least variance of performance across different iterations in both PD and healthy controls (*ps* < 0.001). This effect was confirmed by Levene’s test to account for the violation of normality assumption, *F*_7, 312_ = 11.36, *p* < 0.001. Investigating the features picked by LASSO when the feature pool included both PACs and conventional EEG features, showed that PAC features were the second most picked features after harmonic parameters. Notably, the average beta values assigned to PAC features in LASSO regressions were significantly positive only in PD group *t*(37) = 9.87, *p* < 0.001.

### PAC Informativeness

Subregions of the saliency map corresponding to PACs of delta-beta, theta-alpha, theta-gamma, alpha-beta, alpha-gamma and beta-gamma showed significantly higher average than the overall mean (*ps_bonf* <0.003) and median (*ps_bonf* < 0.045) of the saliency map across all conditions. As a result, in the subsequent analyses, we excluded the delta-theta, delta-alpha, delta-gamma, and theta-beta PACs since their informativeness across the saliency maps were low.

The spatial distribution of the consistency of informativeness for delta-beta, theta-gamma, alpha-gamma, and beta-gamma PACs is shown in Figure 5. The head plots demonstrate the scaled values of the similarity score average for each channel computed within PAC regions on the PACNET saliency maps. These values are controlled for their variability in the TensorPAC inputs. The warmer the color of the channels, the more consistent and informative they were for the PACNET across different trials. Since alpha-beta and theta-alpha PACs showed no difference between their similarity scores in saliency maps and TensorPAC images—their statistical masks were either negative or zero (See Figure 5.B and D)—suggesting that they were not providing consistent information to the PACNET, hence removed from the subsequent analyses.

**Figure 5.**
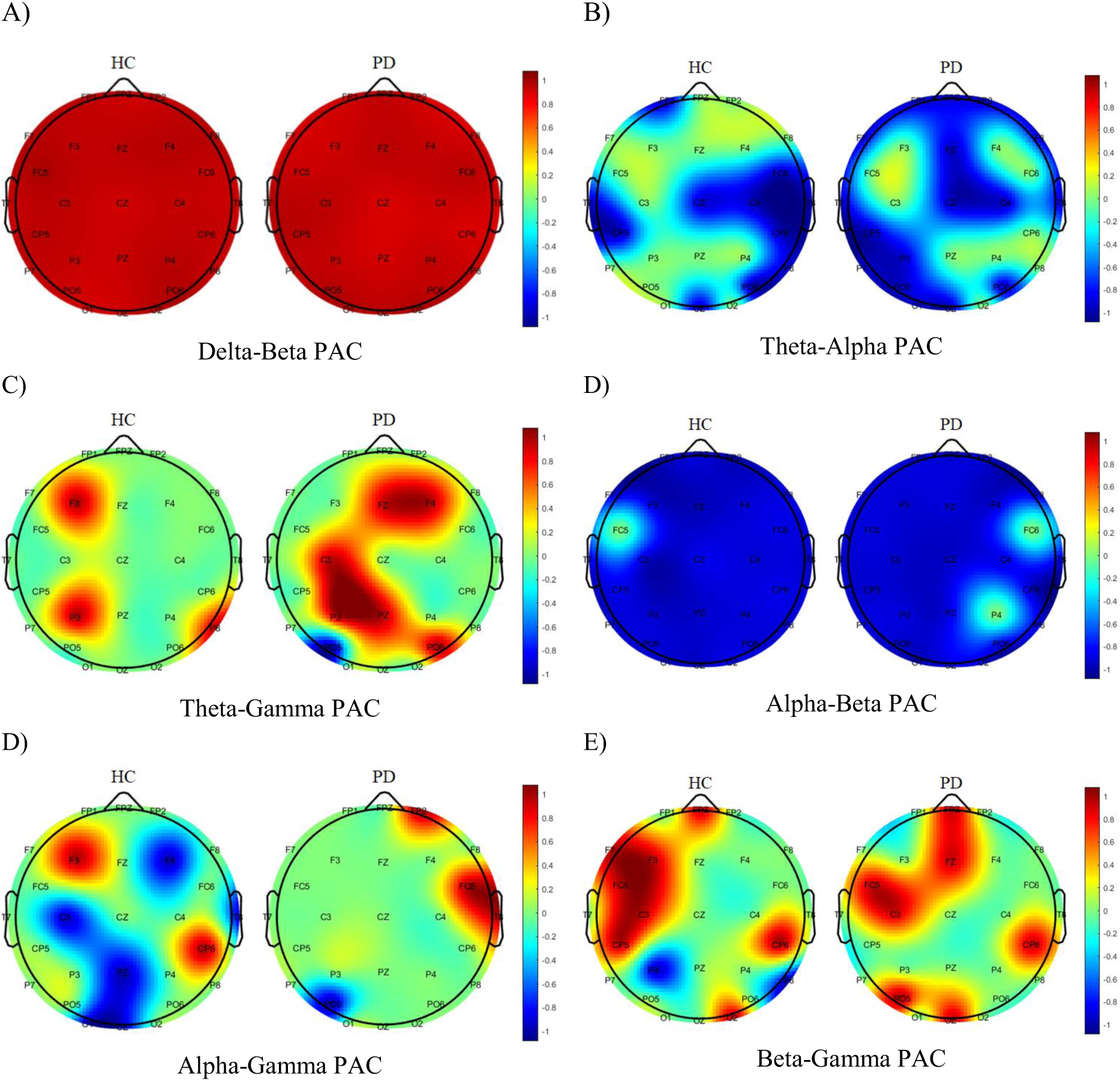
Spatial distribution of the consistency of informativeness for different PACs. Red areas are corresponded to statistically significant consistency of informativeness (p<0.05).

Delta-beta PAC across both HC and PD groups was found to be informative across all channels (Figure 5.A). In HC, MV-associated theta-gamma PAC was predominantly localized over the left frontal (F3), left parietal (P3), and left parieto-occipital (P6) regions. Conversely, in individuals with PD, theta-gamma PAC was observed in different regions, including the frontal midline (Fz), right frontal (F4), left central (C3), and left parietal (P3) areas (Figure 5.C). In HC, alpha-gamma PAC was predominantly localized over the left frontal (F3) and right parietal (CP6) regions. In PD, alpha-gamma PAC was observed in different regions, including the right frontal (FP2) and right central (FC6) areas (Figure 5.D). In HC, beta-gamma PAC (the most important PAC for PD) was predominantly localized over several frontal and central regions, including FPz, F3, FC5, C3, CP5, CP6, and O2. In PD, Beta-gamma PAC was observed in some overlapping regions, including FPz, FC5, C3, CP6 and Oz (Figure 5.E).

To compare the overall informativeness of different PACs, we ran a 2 (Health: HC vs. PD) by 4 (PACs: delta-beta, theta-gamma, alpha-gamma, and beta-gamma) mixed ANOVA on the Z-transformed average of the similarity scores across channels with non-negative consistency of informativeness. We found a main effect of health *F*_1, 78_ = 89.73, *p* < 0.001, such that PACs informativeness was more consistent in PD groups than HC, *t*(78) = -9.47, *p* < 0.001. We also found a significant main effect of PACs, *F*_3, 234_ = 916.33, *p* < 0.001 which was confirmed by a significant interaction between health and PACs, *F*_3, 234_ = 9.32, *p* < 0.001, such that informativeness between PACs from the most consistent to the least one can be ranked as alpha-gamma, theta-gamma, beta-gamma, and delta-beta (See Figure 6).

**Figure 6.**
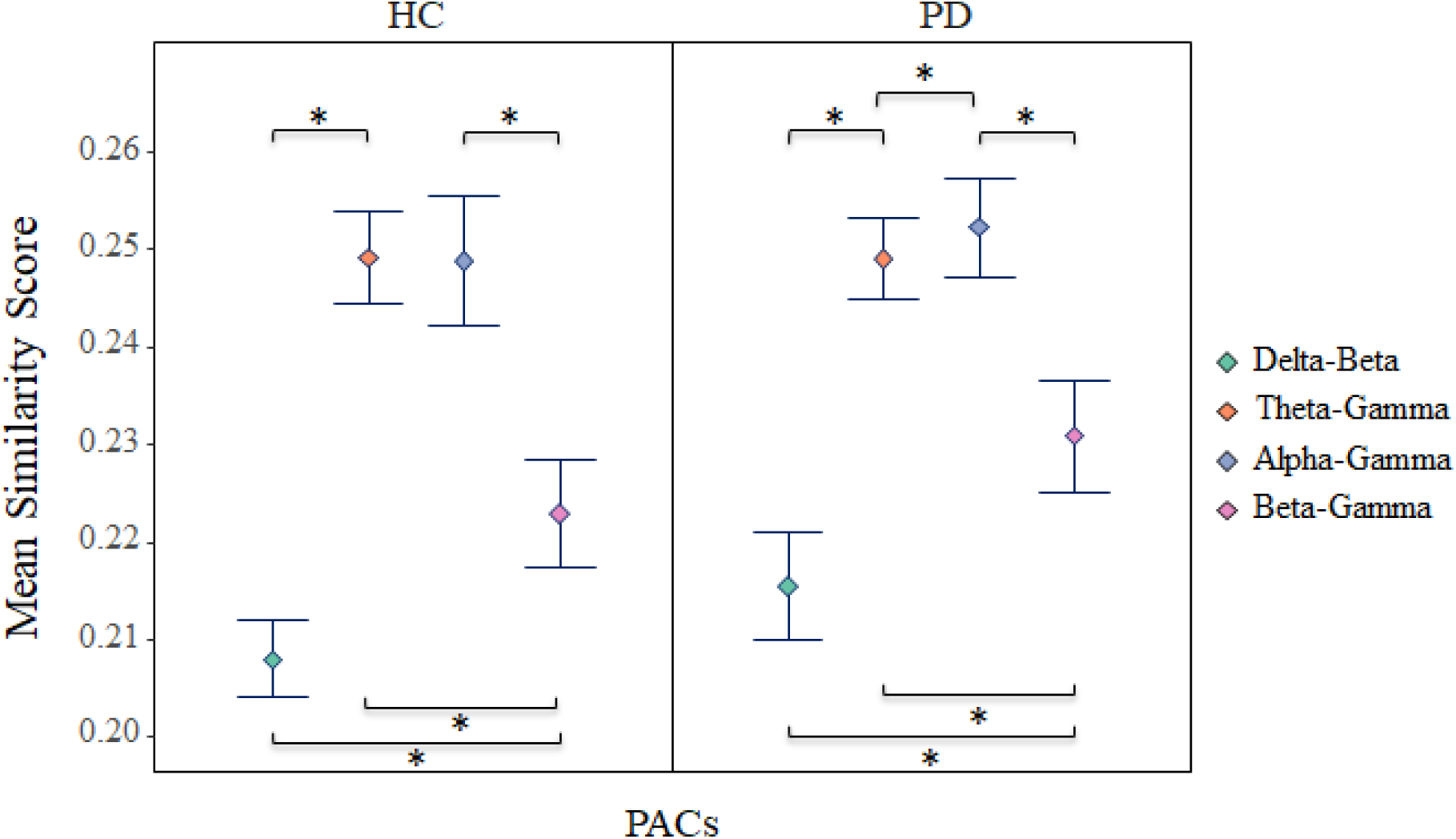
The mean value of the similarity scores in sham and no medication conditions for healthy and PD groups. *indicates that the Bonferroni corrected p-value was significant (<0.05). Error bars indicate the standard deviation around the mean.

### Stimulation and Medication Effects

This analysis aims to explore whether GVS and L-dopa medication influence the specific types of PAC that are most informative for predicting motor vigor (MV). We will also examine whether these effects differ between PD-on, PD-off and healthy controls. Within each PAC on the Fisher Z transformed value of the similarity scores, we conducted a 2 (Health: HC and PD-off) by 3 (Stimulation: Sham, GVS1, and GVS2) mixed ANOVA to investigate the effect of stimulation on the informativeness of each PAC between PD and healthy controls and a 2 (Health: PD-Off and PD-On) by 3 (Stimulation: Sham, GVS1, and GVS2) mixed ANOVA to investigate the effect of stimulation on the informativeness of each PAC within PD groups with or without medication. Figure 7 shows the mean value of the similarity scores of different PACs across the iterations.

**Figure 7.**
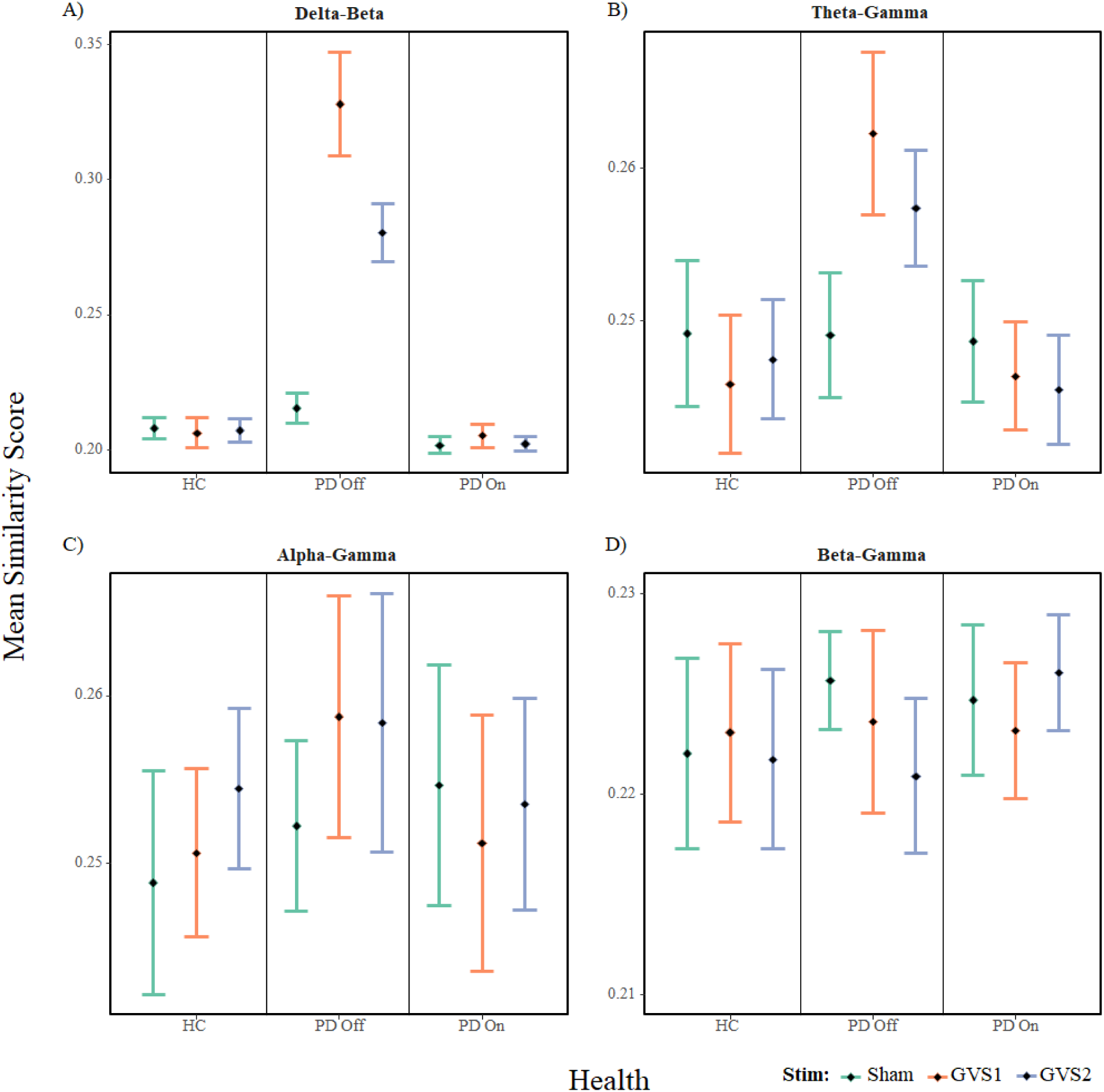
The mean value of the similarity scores across the iterations for each PAC across populations for 3 different Stim conditions. Error bars indicate the standard deviation around the mean.

We observed higher similarity scores for PD subjects off medication than HC across delta-beta, theta-gamma, alpha-gamma, and beta-gamma (*ps* < 0.001). We also found a main effect of L-dopa medication across delta-beta, theta-gamma, alpha-gamma, and beta-gamma (*ps* < 0.034). Within the HC group, alpha-gamma differentiated all three stimulation conditions (*ps* < 0.002), and theta-gamma differentiated between GVS1 and Sham (*p* = 0.002). Within the PD-off group, all stimulation conditions could be reliably dissociated based on delta-beta (*ps* < 0.001), beta-gamma (*ps* < 0.004), and theta-gamma (*ps* < 0.016) PACs while alpha-gamma could only reliably dissociate Sham from GVS1 and GVS2 (*ps* < 0.001). Within the PD-on group, delta-beta was only informative for GVS1 (*ps* < 0.002), while theta-gamma reliably dissociated Sham from both GVS1 and GVS2 (ps < 0.024), and finally, beta-gamma reliably dissociated the GVS1 and GVS2 (*p* = 0.001). Details of the ANOVA tests and their corresponding post-hoc t-tests are provided in the supplementary material section 7.

## Discussion

In this study, we explored the relation between motor vigor, Parkinson’s disease (PD), and electroencephalogram (EEG) signals using a comprehensive analytical approach that involved phase-amplitude coupling (PAC) analysis. Our analysis focused on PACs within different EEG frequency bands, including delta-beta, theta-gamma, alpha-beta, alpha-gamma, and beta-gamma, and their potential informativeness in predicting motor vigor. For this purpose, we employed a deep neural network (DNN) model, PACNET, which allowed us to capture complex, non-linear interactions among PACs. The results of our study offer valuable insights into several critical aspects.

### Interpretation of the Results

Prior works on PAC in PD have focused on rest, but a recent study demonstrated that beta-gamma PAC was altered during transitions between different movement states (Gong et al., 2022). Our results show that PAC, at multiple frequency combinations, is directly related to MV, even in a single, overlearned task which would not necessarily involve switching between movement states. While PAC appears directly related to MV in healthy controls, different PACs are sensitive to the development of PD, the effects of dopaminergic medication and the effects of non-invasive brain stimulation in PD in a stimulus-sensitive manner.

We observed that delta-beta PAC exhibited a strong association with motor vigor in the PD-off group. Prior studies have investigated the relationship between the phase of cortical delta oscillations and reaction time. In one study (Stefanics et al., 2010), an auditory target detection paradigm was used to investigate the effect of expectancy on delta activity in the EEG. The phase of delta oscillations significantly entrained to the target onset preceding stimulus processing, with increasing accuracy at higher expectancy levels. Stefanics et al., (2010) further showed that the reaction times correlated significantly with the delta phase at target onset, with the fastest reactions observed when the delta phase at the target onset fell on the rising slope of delta oscillations. In a rat model, during a two-alternative forced-choice task, neurons in the motor thalamus that were correlated with movement speed were entrained to delta oscillations and were associated with power fluctuations in beta/low gamma bands (Gaidica et al., 2020). It is unclear why delta-beta PAC would be particularly relevant for PD subjects during GVS stimulation (Figure 7). The ventrolateral nuclei of the thalamus (ventral anterior, VA; ventral lateral, VL) project to the primary motor and premotor cortices, suggesting that this circuit may represent a major vestibular motor pathway (Wijesinghe et al., 2015). We speculate the importance of GVS’s effects on delta-beta PAC in PD is related to GVS’s effects on the motor thalamus, as we have previously demonstrated (Lee et al., 2023).

Theta-gamma PAC was important for predicting MV, particularly in the PD-off group. However, theta-gamma PAC is typically associated with working memory, perception, attention, and learning, not MV (Papaioannou et al., 2022). An fMRI study investigating working memory impairment in PD found increased activation in parietal, limbic and cerebellar regions, possibly as a compensatory strategy (Rottschy et al., 2013). Bradykinesia and working memory are correlated in PD (Ekman et al., 2012), likely because they are both part of a dopamine-sensitive fronto-striatal network (Kehagia et al., 2012). This suggests a correlative, as opposed to a causal connection between theta-gamma PAC and MV in PD.

Alpha-gamma PAC, although showing less differentiation between PD and controls, highlighted potential interactions between medication and stimulation in predicting MV, particularly in the GVS1 condition. This suggests that this PAC may be sensitive to the combined effects of medication and non-invasive stimulation. Unsurprisingly, beta-gamma PAC’s association with MV differed between PD-off and HC, indicating its potential as a marker of motor dysfunction in PD. Gamma oscillations appear intricately related to motor vigor, as they appear at movement onset and are distributed around the motor network of the primary motor cortex, the basal ganglia and motor thalamus (Fischer, 2021).

### Clinical Implications

The identification of informative PACs holds promising clinical implications, as they may serve as potential biomarkers for assessing motor function in PD patients, aiding in the diagnosis and monitoring of disease progression. Furthermore, our results suggest that specific PACs may respond differently to medication and stimulation, opening the door to personalized treatment strategies. Tailoring interventions based on individual PAC profiles could enhance the efficacy of therapeutic approaches toward precision stimulus strategy.

In conclusion, our study employs a novel approach to examine the relationship between PACs in EEG signals and motor vigor in Parkinson’s disease. We identified specific PACs, such as delta-beta, theta-gamma, alpha-beta, alpha-gamma, and beta-gamma, demonstrating consistent informativeness in predicting motor function. These findings provide new insights into the neural mechanisms underlying motor dysfunction and offer promising biomarkers for clinical applications.

One of the critical limitations of this work is the relatively small number of subjects. The proposed model will be required to be validated utilizing a richer EEG database that includes information on a larger PD population before it can be made into a helpful Computer-Aided Diagnostics known as CAD tool for PD.

The approaches used in this paper cannot infer causal linkages between PACs and MV, only associations. However, given that PAC waveforms can be integrated into brain stimulation paradigms (Alekseichuk et al., 2016), a potential causal connection could later be explored.

## Supporting information

Sepplementary Material

## Data Availability

All data produced in the present study are available upon reasonable request to the authors

https://github.com/sa-nouri/pacnet-neuro-vigor

## Acknowledgment

This work was supported by resources made available through the Dynamic Brain Circuits cluster and the NeuroImaging and NeuroComputation Centre at the UBC Djavad Mowafaghian Centre for Brain Health (RRID SCR_019086). MJM was supported by the John Nichol Chair in Parkinson’s Research.

## Code/Data Availability

The GitHub repository, including the codes of this paper, can be accessed here: https://github.com/sa-nouri/pacnet-neuro-vigor. All data produced in the present study are available upon request to the authors.

## Author Contributions

AK: Methodology (LASSO, classic features), software, formal analysis, investigation, data curation, writing - review & editing. SN: Methodology (PACNET), software, investigation, formal analysis, writing – original draft. MM: Conceptualization, project administration, supervision, writing - review & editing. SL: Data collection and pre-processing, writing - review & editing. MM (PI): Supervision, resources, funding acquisition, clinical interpretation, writing - review & editing.

