## Supplementary material for "Predicting Motor Vigor from EEG Phase Amplitude Coupling (PAC) in Parkinson’s Disease: Effects of Dopaminergic Medication and Non-invasive Modulation": Sepplementary Material

### Supplementary Materials

#### 1. Overview

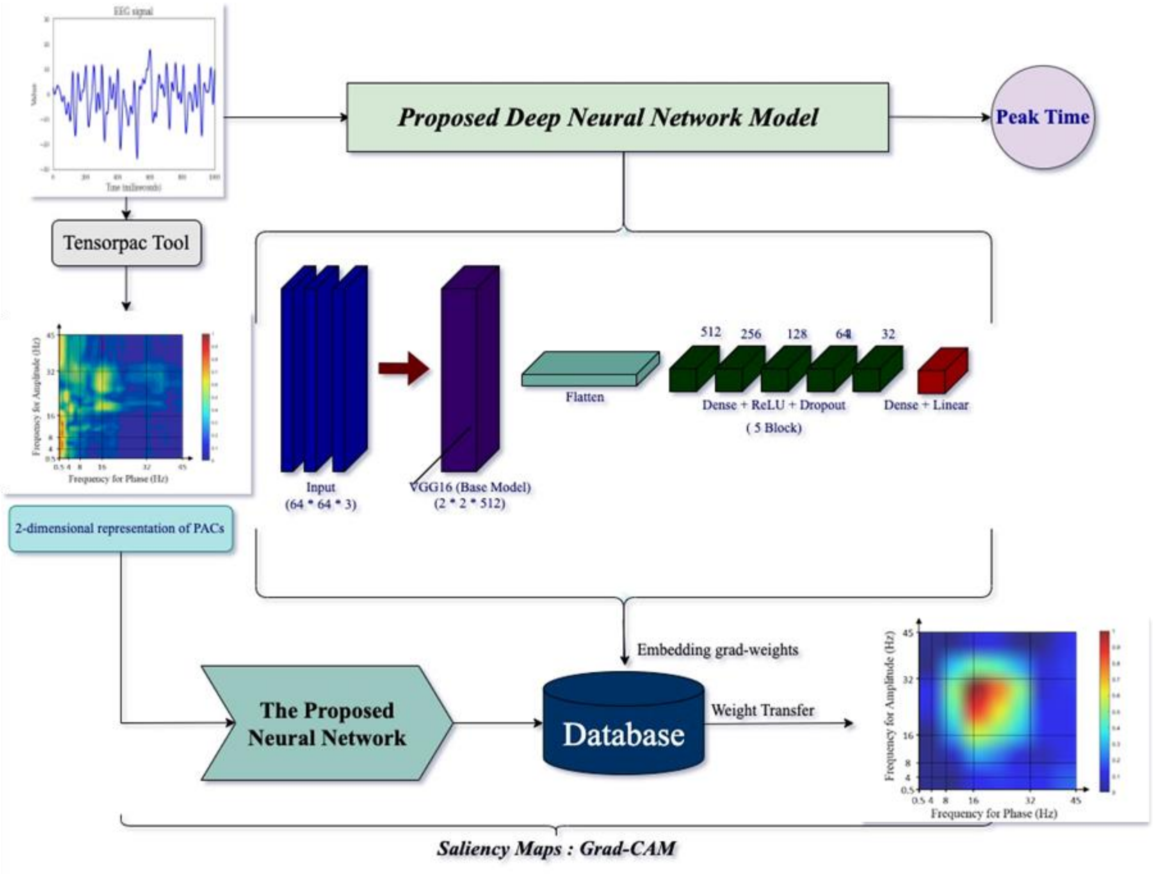


#### 2. Phase-amplitude coupling

Testing for a connection between the phase of one frequency band and the power of another, generally higher frequency band, is what PAC entails. Exploratory approaches for phase and power should be employed to compute experimental phase-amplitude coupling. The quantification of PAC is based on Euler’s formula. PAC uses the time-varying power time series from a higher frequency band. A CFC measure should indicate the relationship between power and simultaneous phase, and the magnitude of power values should not arbitrarily impact it. The PAC value is compared to a distribution of PAC values anticipated under the null hypothesis using non-parametric permutation testing. Permutation testing for PAC entails altering the power time series by a random temporal offset while keeping the phase angle time series unchanged. Under the null hypothesis, the phase-amplitude coupling value is computed using (Canolty et al., 2006):

$$PAC = |\frac{1}{n}\sum_{t=1}^{n} a_t e^{j\phi_t}|. \quad (1)$$

The Observed PAC values are compared to the distribution of PAC values under the null hypothesis by subtracting the mean and dividing by the standard deviation, creating a standardized Z-value of PAC (PACz). Normal-Z values are independent of the scale of the original data, are unaffected by violations of a von Mises distribution, and the result is not affected by substantial power fluctuations.

The value represents the PAC between the respective phase and power frequencies at each position in the two-dimensional (2-D) matrix. In addition, PAC is a metric for detecting and quantifying phase synchronization between low-frequency and high-frequency oscillations. We have employed phase-locking value (PLV) (Penny et al., 2008) for PAC estimation for each pair of angle and power frequencies to create the 2-D matrix because the exaggerated coupling between β-phase and broadband γ-amplitude characterizes PD (de Hemptinne et al., 2013) and the coupling between different neural networks in multiple brain areas engaged in motor control leads to enhanced PAC in PD. The coupling of beta and gamma signals from separate regions seems pathophysiological (Gong et al., 2021). PLV solely considers phase consistency across trials. The PLV is computed by first extracting the phase of the amplitude ϕ, then deducting it from the phase of slower oscillations, then mapping the resulting time series onto the sophisticated circle, and lastly computing the mean of the length vector:

$$PLV = \frac{1}{n}|\sum_{k=1}^{n} e^{j(\phi(k)-\phi_a(k))}|. \quad (2)$$

### 3. Model Architecture

When the feature space distribution alters, the conventional machine-learning algorithms must be recreated from scratch. However, transfer learning transcends the separated learning framework and applies knowledge obtained for one task to tackle relevant ones (Tan et al., 2018). Convolutional Neural Network (CNN) models are acknowledged for their image processing capabilities because of their perfect performance on the ImageNet Large Scale Visual Recognition model; hence, to retrieve high-level features, their primary layers might be employed (Donahue et al., 2014).

**Table 2.** Architecture and parameters of the proposed CNN model

| Layer | Value | Output Shape | Number of parameters |
|---|---|---|---|
| Input | - | (32, 64, 3) | 0 |
| Functional VGG16 | 32 | (1, 2, 512) | 14714688 |
| Flatten | - | (1024) | 0 |
| Batch Normalization | - | (1024) | 4096 |
| Dense + ReLU | - | (4096) | 4198400 |
| Dropout | 0.3 | (4096) | 0 |
| Dense + ReLU | - | (1024) | 4195328 |
| Dropout | 0.3 | (1024) | 0 |
| Dense + ReLU | - | (512) | 524800 |
| Dropout | 0.3 | (512) | 0 |
| Dense + ReLU | - | (256) | 131328 |
| Dropout | 0.3 | (256) | 0 |
| Dense + ReLU | - | (128) | 32896 |
| Dropout | 0.3 | (128) | 0 |
| Dense + ReLU | - | (64) | 8256 |
| Dropout | 0.3 | (64) | 0 |
| Dense + Linear | 1 | (1) | 65 |

| Trainable parameters | Non-trainable parameters | Total parameters |
|---|---|---|
| 9,093,121 | 14,716,736 | 23,809,857 |

The proposed model comprises functional VGG-16, linear, and dropout layers. Convolutional layers of the VGG-16 convolve several kernels with input images to create various feature patterns (Simonyan & Zisserman, 2015). Batch normalization is a method used to normalize inputs to a network. It may be applied to either the activation functions of a preceding layer or inputs directly. It also speeds up training and provides some regularization, which reduces generalization errors (Ioffe & Szegedy, 2015).

Subsequently, a flattened layer is employed, which converts the feature maps into a 1-dimensional feature vector as an input into the next layer. Nodes in the fully connected layers are neurons trained to identify and estimate a single feature vector. The dimension gradually decreased after VGG-16 block processing and flattened into one feature vector of 2048 length. A dropout is a regularization approach that approximates training many neural networks with various designs simultaneously to reduce overfitting and improve generalization error(Srivastava et al., 2014). The neurons in the fully connected layer are trained using the produced single feature vectors. The proposed model's end layer is a linear regression, which regresses the MV by estimating an output value.

### 4. Performance metrics

The evaluation metrics comprise Root Mean Square Error (RMSE), Mean Absolute Error (MAE), and R2-score as Mean Correlation (MC). The RMSE is a quadratic scoring method that determines the error's average size. It's the square root of the average squared differences between predicted and observed values. MAE is a statistic that quantifies the average magnitude of mistakes in a set of predictions without considering their direction. It's the average of the absolute differences between prediction and actual observation over the test sample, where all individual differences have equal weight. R denotes the proportion of variation in the output-dependent attribute that the input-independent variable can predict. It is also used to determine how effectively the model captures observed results based on the ratio of total deviation of results explained by the model. The formulas for each referred metric are described below:

$$MAE = \frac{1}{N}\sum_{j=1}^{N}|y_j - \hat{y}_j| \ , \tag{3}$$

$$RMSE = \sqrt{\frac{1}{N}\sum_{j=1}^{N}(y_j - \hat{y}_j)^2} \ , \tag{4}$$

$$R2 - score = 1 - \frac{SS_{residual}}{SS_{total}} = 1 - \frac{\sum_{j=1}^{N}(y_j - \hat{y}_j)^2}{\sum_{j=1}^{N}(y_j - \bar{y}_j)^2} \tag{5}$$

Where y, yˆ, y¯, and N are the actual value, predicted value, mean value of the variable/feature, and the number of samples, respectively. The model's performance is assessed using particular 40-fold cross-validation, in which specific trials of each participant's data are selected as the test data. The remaining are training and validation data (80 percent training dataset and 20 percent validation dataset). The model will be trained using the training dataset and validation dataset to optimize the model, and the model's performance will be evaluated using the selected test data. The performance of the suggested model in terms of the loss function and metrics is high for training, validation, and test datasets.

### 5. Visual explanation approaches

The proposed model's learned weights were saved to assess the features' representations in each data sample and determine clinical significance. A reasonable visual explanation method that can be used to explain any target (MV value and conditions) should have two properties: class discriminability and high resolution. It allows us to localize different regions in the input image, contributing to other output values and capturing fine-grained detail.

Gradient-weighted Class Activation Mapping (Grad-CAM) uses gradients of any target concept flowing into the final convolutional layer to produce a coarse localization map highlighting the important regions in the image for estimating the idea (Selvaraju et al., 2017). Deep layers in a CNN capture higher-level visual constructs. Furthermore, convolutional layers naturally retain spatial information, which is lost in fully connected layers. The last convolutional layers have the best compromise between high-level semantics and detailed spatial information. We aim to get the features detected in the last convolution layer and examine which ones are the most active for high/low motor vigor values. To do so, each channel gets multiplied in the feature map array of the last convolutional layer about the output class and then sums all the channels to obtain a heatmap of relevant regions in the image (see the supplementary materials section 6 for more details). The backpropagation visualization (saliency map) (Simonyan et al., 2013) can be generated by getting the gradient of the loss concerning the image pixels. The changes in specific pixels that strongly affect the loss will be shown brightly. However, this often produces a noisy image, and it has been demonstrated that clipping the gradients less than zero during backpropagation (intuitively allowing only positive influences) gives a sharper image. The Guided Backpropagation technique produces a heatmap of gradients of the same size as the input image. Since we do not need to resize a small feature map to the size of the input image, this is by high construction resolution. However, the drawback here is that the visualization is not class discriminative.

### 6. Details of Statistical Tests

According to fitting/training parameters, the deep Model converged on the data appropriately. A one-way repeated measure analysis of variance (ANOVA) is utilized to determine whether there are any statistically significant differences among the PAC values of each condition with others. The aim of this evaluation is that the model's inputs are significantly different from each other, and the PAC values engulf the pertinent information about the conditions, such as stimuli, healthy, and PD.

| Study cases for PAC | Sham vs. Stim7 | Sham vs. Stim8 | Stim7 vs. Stim8 |
|---|---|---|---|
| P-value | 4.62 e-7 | 6.37 e-5 | 5.82 e-8 |

| Study cases for PAC | HC vs. PD-off | HC vs. PD-on | PD-on vs. PD-off |
|---|---|---|---|
| P-value | 3.48 e-5 | 7.52 e-4 | 6.74 e-5 |

The tables above indicate that these study cases are significantly different from each other. So, the input of the proposed model has the necessary information for training purposes. Moreover, the absolute error of the predicted and actual MV values for each data is calculated after training the model. R-2 score also is computed for predicted MV values in each fold for further statistical tests. Then, one-way repeated measures ANOVA is employed to test the significant differences of these performance metrics for each condition. The table below illustrates the p-value of the tests.

| Study cases for MAE | Sham vs. Stim7 | Sham vs. Stim8 | Stim7 vs. Stim8 |
|---|---|---|---|
| P-value | 0.731 | 0.657 | 0.782 |

| Study cases for MAE | HC vs. PD-off | HC vs. PD-on | PD-on vs. PD-off |
|---|---|---|---|
| P-value | 0.252 | 0.315 | 0.382 |

| Study cases for Correlation | Sham vs. Stim7 | Sham vs. Stim8 | Stim7 vs. Stim8 |
|---|---|---|---|
| P-value | 0.375 | 0.431 | 0.369 |

| Study cases for Correlation | HC vs. PD-off | HC vs. PD-on | PD-on vs. PD-off |
|---|---|---|---|
| P-value | 0.865 | 0.458 | 0.641 |

The tables above denote that the trained model gives the same level of performance across groups, which allows us to use the learned knowledge of the model for further exploration, for instance, exploring saliency maps to find influential PACs for PD due to the proof of PAC informativity.

### 7. Details of the Statistical tests for the Health and Medication Effects

The average and standard deviations reported here are average of fisher Z transformed values of the similarity scores. Within **Delta-Beta PAC** between HC and PD, we found a significant interaction effect between health and stimulation *f*(2, 156) = 633.75, $p<0.001$, $\eta_p^2 = .89$ which is confirmed by significant main effect in both health condition, *f*(1, 78) = 2836.78, $p<0.001$, $\eta_p^2 = .97$, and stimulation *f*(2, 156) = 596.98, $p<0.001$, $\eta_p^2 = .88$. Critically, similarity scores for PD Off, (*M* = 0.28, *SD* = 0.01), were significantly higher than HC, (*M* = 0.21, *SD* < 0.01); *t*(78) = 53.26, $p < 0.001$, Cohen's *d* = 11.91, *pbonf.* < 0.001, *95% CI* [0.07, 0.07], which suggests that Delta-Beta was generally a more informative feature for the PD-off group. Delta-Beta did not capture the effect of stimulation in HC group (*ps* > 0.056, *pbonf.* > 0.168) while in PD-off group, GVS1, (*M* = 0.34, *SD* = 0.02) were significantly higher than GVS2, (*M* = 0.29, *SD* = 0.01); *t*(39) = 12.62, $p < 0.001$, Cohen's *d* = 2, *pbonf.* < 0.001, *95% CI* [0.04, 0.06] which in turn were significantly higher than Sham, (*M* = 0.22, *SD* = 0.01); *t*(39) = 32.51, $p < 0.001$, Cohen's *d* = 5.14, *pbonf.* < 0.001, *95% CI* [0.06, 0.07] (See Figure 7.A). Similarly, between PD-off and PD-on, we found a significant interaction effect between medication and stimulation *f*(2, 78) = 610.13, $p < 0.001$, $\eta_p^2 = .94$ which was confirmed by significant main effect in both medication condition, *f*(1, 39) = 3923.89, $p < 0.001$, $\eta_p^2 = .99$, and stimulation *f*(2, 78) = 655.18, $p < 0.001$, $\eta_p^2 = .94$. Similarity scores for PD-off, were significantly higher than PD-on, (*M* = 0.21, *SD* < .01); *t*(39) = 62.64, $p < 0.001$, Cohen's *d* = 9.9, *pbonf.* < 0.001, *95% CI* [0.07, 0.08], which suggests that medication effect on Delta-Beta was generally a more informative feature for the PD-off group. Delta-Beta captured the effect of stimulation in PD-on group as well such that GVS1, (*M* = 0.208, *SD* < .01) were significantly higher than both Sham, (*M* = 0.205, *SD* < .01); *t*(39) = 4.24, $p < 0.001$, Cohen's *d* = 0.67, *pbonf.* < 0.001, *95% CI* [.002, .005] and GVS2, (*M* = 0.205, *SD* < .01); *t*(39) = 3.8, $p = 0.001$, Cohen's *d* = 0.6, *pbonf.* = 0.002, *95% CI* [.001, .005] (See Figure 7.A). Overall, Delta-Beta PAC could successfully capture both medication and stimulation effects on PD groups and dissociate

PD from HC. This PAC involves the coupling between delta (slow) and beta (relatively faster) brain oscillations.

Within **Theta-Gamma PAC** (See Figure 7b) between HC and PD, we found a significant interaction effect between health and stimulation *f*(2, 156) = 76.79, $p < 0.001$, $\eta_p^2 = .05$ which was confirmed by significant main effect in both health, *f*(1, 78) = 195.98, $p < 0.001$, $\eta_p^2 = .72$, and stimulation *f*(2, 156) = 28.51, $p < 0.001$, $\eta_p^2 = .27$. Similarity scores for PD-off (*M* = 0.26, *SD* = 0.01), were significantly higher than HC, (*M* = 0.25, *SD* = 0.01); *t*(78) = 14, $p < 0.001$, Cohen's *d* = 3.13, *pbonf.* < 0.001, *95% CI* [.008, .01], which suggests that Theta-Gamma was generally a more informative feature for the PD-off group. Within PD-off group, Theta-Gamma PAC similarity scores were significantly higher for GVS1, (*M* = 0.27, *SD* = 0.01) compared to GVS2, (*M* = 0.26, *SD* < 0.01); *t*(39) = 4.67, $p < 0.001$, Cohen's *d* = 0.739, *pbonf.* < 0.001, *95% CI* [0.003, 0.01] which in turn was higher than Sham, (*M* = 0.25, *SD* < 0.01); *t*(39) = 10.03, $p < 0.001$, Cohen's *d* = 1.59, *pbonf.* = 0.016, *95% CI* [.007, .01]. Within HC group, Sham (*M* = 0.255, *SD* = 0.01), were significantly higher than GVS1, (*M* = 0.251, *SD* = 0.01); *t*(39) = 3.67, $p = 0.001$, Cohen's *d* = 0.58, *pbonf.* = 0.002, *95% CI* [.002, 0.01]. Between PD-off and PD-on, there was a significant interaction between medication and stimulation, *f*(2, 78) = 67.94, $p < 0.001$, $\eta_p^2 = 0.64$ which was confirmed by a significant main effect in both medication *f*(1, 39) = 280.7, $p < 0.001$, $\eta_p^2 = 0.88$; and stimulation, *f*(2, 78) = 49.89, $p < 0.001$, $\eta_p^2 = 0.56$, such that similarity scores for PD-off, (*M* = 0.26, *SD* < 0.01), were significantly higher than PD-on, (*M* = 0.25, *SD* < 0.01); *t*(39) = 16.78, $p < 0.001$, Cohen's *d* = 2.65, *pbonf.* < 0.001, *95% CI* [0.01, 0.01]. Importantly, theta-gamma captured the effect of stimulation in PD-on group such that Sham, (*M* = 0.254, *SD* < 0.01), were significantly higher than GVS1, (*M* = 0.252, *SD* < 0.01); *t*(39) = 2.79, $p = 0.008$, Cohen's *d* = 0.44, *pbonf.* =

0.024, *95% CI* [0.001, 0.004], and GVS2, (*M* = 0.251, *SD* < 0.01); *t*(39) = 4.22, *p* < 0.001, Cohen's *d* = 0.67, *pbonf.* < 0.001, *95% CI* [0.002, 0.01].

Within **Alpha-Gamma PAC** (See Figure 7c) between HC and PD, we found a significant interaction between health and stimulation, *f*(2, 156) = 3.61, *p* = 0.03, $\eta_p^2$ = 0.04 which was confirmed by main effects of health, *f*(1, 78) = 39.53, *p* < 0.001, $\eta_p^2$ = 0.34 and stimulation *f*(2, 156) = 19.52, *p* < 0.001, $\eta_p^2$ = 0.2. Similarity scores for PD-off (*M* = 0.262, *SD* < 0.01), were significantly higher than HC, (*M* = 0.257, *SD* < 0.01); *t*(78) = 6.287, *p* < 0.001, Cohen's *d* = 1.41, *pbonf.* < 0.001, *95% CI* [.004, 0.01], which suggests that alpha-gamma was generally a more informative feature for the PD-off group. Within the PD-off group, alpha-gamma PAC could dissociate sham, (*M* = 0.258, *SD* = 0.01) from both GVS1, (*M* = 0.265, *SD* = 0.01); *t*(39) = -4.4, *p* < 0.001, Cohen's *d* = -0.7, *pbonf* < 0.001, *95% CI* [-0.01, -0.004] and GVS2, (*M* = 0.264, *SD* = 0.01); *t*(39) = -4.23, *p* < 0.001, Cohen's *d* = -0.67, *pbonf* < 0.001, *95% CI* [-0.01, -0.003]. Within HC group, GVS2, (*M* = 0.26, *SD* = 0.01) were significantly higher than both GVS1, (*M* = 0.256, *SD* = 0.01), *t*(39) = 3.68, *p* = 0.001, Cohen's *d* = 0.58, *pbonf.* = 0.002, *95% CI* [0.002, 0.006] and Sham, (*M* = 0.254, *SD* = 0.01), *t*(39) = 4.372, *p* < 0.001, Cohen's *d* = 0.69, *pbonf.* < 0.001, *95% CI* [0.003, 0.009]. Critically, in the PD-off group, alpha-gamma showed consistency of informativeness in the same level for both GVS1 and GVS2 and higher than Sham. Between PD-off and PD-on, there was a significant interaction between medication and stimulation, *f*(2, 78) = 13.11, *p* < 0.001, $\eta_p^2$ = 0.25 and a significant main effect of medication, *f*(1, 39) = 13.62, *p* < 0.001, $\eta_p^2$ = 0.26, such that similarity scores for PD-off, (*M* = 0.262, *SD* < 0.01), were significantly higher than PD-on, (*M* = 0.259, *SD* < 0.01); *t*(39) = 3.69, *p* = 0.001, Cohen's *d* = .58, *95% CI* [0, 0.01].

Within **Beta-Gamma PAC** (See Figure 7d) between HC and PD-off, we found a significant interaction effect between health and stimulation *f*(2, 156) = 15.77, *p* < 0.001, $\eta_p^2$ = .17

which was confirmed by significant main effect in both health condition, $f(1, 78) = 16.35$, $p < 0.001$, $\eta_p^2 = .17$, and stimulation $f(2, 156) = 13.2$, $p < 0.001$, $\eta_p^2 = .14$. Similarity scores for PD-off ($M = 0.23$, $SD < 0.01$), were significantly higher than HC, ($M = 0.227$, $SD < 0.01$); $t(78) = 4.04$, $p < 0.001$, Cohen's $d = 0.9$, *pbonf.* < 0.001, *95% CI* [.001, 0.004], which suggests that Beta-Gamma was generally a more informative feature for the PD off group. Beta-Gamma PAC did not capture the effect of stimulation in HC group (*ps*>0.674) while in PD-off group, could dissociate all stimulations consistently such that similarity scores for Sham, ($M = 0.24$, $SD = 0.01$), were significantly higher than GVS1, ($M = 0.23$, $SD = 0.01$); $t(39) = .643$, $p < 0.001$, Cohen's $d = 0.64$, *pbonf.* = 0.001, *95% CI* [0.003, 0.01] which in turn was significantly higher than GVS2, ($M = 0.225$, $SD = 0.01$); $t(39) = 3.46$, $p = 0.001$, Cohen's $d = 0.55$, *pbonf.* = 0.004, *95% CI* [0.002, 0.01]. Critically, between PD-off and PD-on, there was a significant interaction between medication and stimulation, $f(2, 78) = 23.83$, $p < 0.001$, $\eta_p^2 = 0.38$ which was confirm by main effect in both stimulation, $f(2, 78) = 12.48$, $p < 0.001$, $\eta_p^2 = 0.24$, and medication, $f(1, 39) = 4.82$, $p = 0.03$, $\eta_p^2 = 0.11$ such that Similarity scores in PD-off, ($M = 0.23$, $SD < 0.01$), were significantly different from PD-on, ($M = 0.232$, $SD < 0.01$); $t(39) = -2.2$, $p = 0.034$, Cohen's $d = -0.35$, *95% CI* [0, 0]. Within PD-on group, GVS1, ($M = 0.229$, $SD = 0.01$), were significantly different from GVS2, ($M = 0.2341$, $SD = 0.01$); $t(39) = -4.021$, $p < 0.001$, Cohen's $d = -0.636$, *pbonf.* = 0.001, *95% CI* [-0.007, -0.002].
